# Oculocutaneous albinism variants in 28 consanguineous families and functional classification of a pathogenic deep intron variant in *TYR*

**DOI:** 10.1101/2025.02.20.25322192

**Authors:** Muhammad Farooq, Gitte Hoffmann Bruun, Menachem V. K. Sarusie, Line Kessel, Hamna Akhtar, Uzma Abdullah, Iram Anjum, Thomas K. Doktor, Brage Storstein Andresen, Shahid Mahmood Baig, Lars Allan Larsen, Karen Grønskov

**Author notes:** Authors contributed equally (Muhammad Farooq, Gitte Hoffmann Bruun). Corresponding authors: Lars Allan Larsen, Department of Cellular and Molecular Medicine, University of Copenhagen, 2200 Copenhagen, Denmark., Karen Grønskov, Department of Genetics, Righospitalet, Copenhagen University Hospital, 2600 Glostrup, Denmark.

## Abstract

Oculocutaneous albinism (OCA) is genetically and clinically heterogeneous recessive disorders with at least 23 associated genes. Isolated OCA is characterized by hypopigmentation in the skin, hair, and eyes combined with ocular abnormalities. Hermansky Pudlak syndrome (HPS) and Chediak-Higaski syndrome are syndromic forms of OCA, distinguished by immunological and hematological symptoms in addition to hypopigmentation and ocular anomalies. Targeted clinical care is crucial for the patients and molecular genetic diagnosis is important for classification of patients. Current diagnostic yield is approximately 70%, and a high prevalence of patients, heterozygous for pathogenic variants in OCA genes, might suggest presence of disease-causing non-coding variants.

We describe here NGS analysis, including CNV analysis, of 28 consanguineous families, comprising a total of 136 individuals presenting with OCA. We provide a molecular genetic diagnosis in all 28 families. Noteworthy, five families (18 %) had pathogenic variants in a gene associated with HPS. Furthermore, we report the first deep intron variant in *TYR* causing OCA and show by minigene analysis that the variant causes inclusion of a pseudoexon.

## Introduction

Oculocutaneous albinism (OCA) is a clinical and molecular heterogenous disorder, with autosomal recessive inheritance (Grønskov et al. 2007; Thomas et al. 1993). Eight distinct clinical types of isolated OCA are known OCA1 (*TYR*), OCA2 (*OCA2*), OCA3 (*TYRP1*), OCA4 (*SLC45A2*), OCA5 (gene unknown), OCA6 (*SLC24A5*), OCA7 (*LRMDA*) and OCA8 (*DCT*). Isolated OCA is characterized by hypopigmentation of skin, hair, and eyes in addition to ocular abnormalities, which include nystagmus, fovea hypoplasia, iris transillumination, photophobia, misrouting of the optic nerves and decreased visual acuity.

In addition, there are syndromic forms of OCA; Hermansky Pudlak syndrome (HPS) and Chediak-Higaski syndrome (CHS). The syndromic forms are characterized by immunological and hematological symptoms in addition to hypopigmentation and eye symptoms. HPS is a genetically heterogeneous disorder caused by defects in function or transport of lysosome related organelles (LRO) and is characterized by hemorrhagic diathesis and OCA, plus other severe clinical symptoms, such as immunodeficiency, pulmonary fibrosis and granulomatous colitis depending on the specific type of HPS. The prevalence of HPS is estimated to be 1/100,000 in most populations, although some populations such as the Puerto Rician has a significantly higher incidence of 1 in 1800 (Wildenberg et al. 1995).

Molecular genetic diagnosis of individuals with albinism is therefore important to identify those with syndromic forms so that accurate clinical care can be undertaken. Even though all genes currently known to be associated with OCA are analyzed, approximately 30% of patients are left without a genetic diagnosis (Chan et al. 2023; Lasseaux et al. 2018). A larger number than expected from the carrier frequency is heterozygous for a pathogenic variant in *TYR*, suggesting that intronic *TYR* variants might play a role in some cases (Grønskov et al. 2009).

Intronic sequence variations may be pathogenic by activating pseudoexons (Petersen et al. 2022). Pseudoexons are exon-like sequences, which are normally not included in the final mRNA transcript, or at least only to a very low degree due to their location in a suboptimal splicing context (Petersen et al. 2022). Sequence variants may strengthen the splicing context of a pseudoexon, for instance by increasing splice site strength, creating a splicing enhancer or by abolishing a splicing silencer, leading to activation and increased inclusion of the pseudoexon. Inclusion of pseudoexons will disrupt the open reading frame, in many cases leading to introduction of premature termination codons (PTC) resulting in a truncated protein and/or often lead to degradation of the resulting mRNA transcript by the nonsense mediated decay (NMD) mechanism. In the present study we analyzed 28 consanguineous OCA families from Pakistan. Genetic diagnosis identified 23 families with isolated OCA and five families with HPS. In one of the non-syndromic OCA families, we identified a very rare deep intron variant in *TYR*, which was shown by minigene analysis to cause inclusion of a pseudoexon. This is the first reported pathogenic deep intron variant in *TYR*.

## Materials and methods

### Consanguineous OCA families

Twenty-eight consanguineous OCA families were identified in the Punjab region of Pakistan based on the presence of hypopigmentation of skin and hair in multiple affected individuals. We identified between 1 and 15 affected individuals per family (median=4), comprising a total of 136 affected (85 males, 52 females). Pedigrees are available on request (see **Fig. S1)**. The study followed the declaration of Helsinki and was approved by the Institutional Review Board of GC University Faisalabad, Faisalabad, Pakistan and Institutional Research Board (IRB), Health Services Academy (HSA), Islamabad, Pakistan. Informed consent was obtained from all participating individuals or their parents for the collection of blood samples, genetic analyses, and publication of genetic information. Blood samples were drawn from affected and unaffected individuals. DNA was extracted using standard methods.

### Molecular genetic analysis

#### NGS gene panel analysis

Targeted Next generation sequencing (NGS) was performed using a SureSelect custom library (Agilent, Santa Clara, California, United States) of a gene panel with 20 genes (*AP3B1*, *BLOC1S3*, *BLOC1S6*, *LRMDA*, *DTNBP1*, *GPR143*, *HPS1*, *HPS3*, *HPS4*, *HPS5*, *HPS6*, *LYST*, *MLPH*, *MYO5A*, *OCA2*, *RAB27A*, *SLC24A5*, *SLC45A2*, *TYR*, *TYRP1*) plus a deep intron variant in *TYR* (rs147546939) known to be associated with a pathogenic haplotype (Grønskov et al. 2019). Three newly identified genes, *AP3D1* (Mohammed et al. 2019), *BLOC1S5* (Pennamen et al. 2020) and *DCT* (Pennamen et al. 2021), were not analyzed as they were identified after the design of the NGS panel. Sequencing was performed using Illumina technology (Illumina, San Diego, California, United States) and MiSeq platform (Illumina). For alignment and variant calling SureCall v. 3.5.1.46 (Agilent) were used with default settings. VarSeq v2.2.3 (Golden Helix, Bozeman, Montana, United States) was used for annotation and filtering of variants. Variants were filtered for MAF < 1%, location in coding exons plus 20 bp of intron sequence adjacent to exons. CNV analysis was performed using the CNV caller in VarSeq. Variants were classified using the ACMG/AMP guidelines (25741868, 30192042, 31892348, 27236918, 30311383, PM2 v 1.0, PS2/PM6 v1.1, PM3 v 1.0, webpage https://clinicalgenome.org/working-groups/sequence-variant-interpretation/).

#### Whole genome sequencing and homozygosity mapping

Samples from a five generation family with six individuals affected by OCA were analyzed by whole genome sequencing. Blood samples were obtained for III.1, III.2, IV.1, IV.2, IV.6 and V.1 and high molecular DNA was extracted using the Chemagic 360 machine (Perkin Elmer, Waltham, Massachusetts, USA). Whole genome sequencing was performed by BGI Group (Shenzhen, China) using Illumina HiSeq X-ten. A mean coverage of 30 X was provided. Alignment was performed to NCBI hg19 version of the human genome using BWA software. GATK (Broad Institute, MIT Harvard, Cambridge, MA, USA) was used for variant calling (SNV and indels). Data was analyzed using VarSeq software v2.2.3 (Golden Helix, Bozeman, Montana) with settings Variant Allele Fraction (VAF) > 0.2, Minor Allele Frequency (MAF) < 0.005 and location in *TYR* both coding and noncoding regions. CNV analysis was performed using the CNV caller in VarSeq.

Homozygozity-by-descent regions spanning more than 1 Mbp in size were identified using the UCSC genome browser. Bed files with colour-coded homozygous and heterozygous variants were created from vcf files.

In silico analysis of splice-site variants were performed using SpliceAI (de Sainte Agathe et al. 2023), Pangolin (Zeng and Li 2022) and MaxEnt Scan (http://hollywood.mit.edu/burgelab/maxent/Xmaxentscan_scoreseq.html). Missense variant effects were analyzed using Combined Annotation Dependent Depletion, CADD (Kircher et al. 2014) and REVEL (Ioannidis et al. 2016).

### Minigene analysis

Minigenes in pcDNA3.1 containing 24 bp of *TYR* exon 4, a reduced intron 4 (the first 480 bp of intron 4, 2520 bp is reduced, 2040 bp intron with an introduction of an EcoRI restriction site after 1017 bp, 4860 deletion, and the last 288 bp of intron 4) and 43 bp of exon 5 carrying either the wild-type or the mutant (NM_000372.4:c.1366+4629A>G) sequence were synthesized by Synbio Technologies (Monmouth Junction, NJ, USA). Minigenes were transfected into RPE-1 cells using X-tremeGENE™ 9 DNA Transfection Reagent (Roche, Basel, Switzerland) following manufacturer’s instructions. RNA was extracted using TRIzol™ Reagent (Thermo Fisher Scientific, Waltham, MA, USA) and was reverse transcribed and subject to PCR analysis using primers forward: GCTGGCTAGCACTATAGCTA and reverse: CCTCTAGACTCGAGTGTTCC. PCR products were separated on 2% agarose gels, and bands were excised and Sanger sequenced by Eurofins Genomics (Ebersberg, Germany).

Splice switching oligonucleotides (SSOs) were synthesized by LGC Biosearch Technologies (Middlesex, UK). The SSOs contained phosphorothioate and 2’OMe-modified backbones with the following sequences: PE2 5’ss: UAAAUACCUUCAAGCUUCUAAUAUG and PE3 5’ss: UACCUUUUCACCCUCAGCACCUGGC. Control SSO: CAAUAUGCUACUGCCAUGCUUG. SSOs were transfected 24 hours after minigenes into RPE-1 cells by Lipofectamine™ RNAiMAX Transfection Reagent (Thermo Fisher Scientific, Waltham, MA, USA).

## Results

### NGS gene panel analysis

Gene panel analysis identified pathogenic variants in 27 families, and whole genome sequencing identified a deep intron variant in *TYR* in one family (**Table 1**). *TYR* was the most prevalent gene explaining OCA in 15 out of 28 (54%) families, followed by *OCA2* which explained 7 out of 28 (25%) of families. All variants were homozygous in affected individuals. A pathogenic variant c.1255G>A in *TYR* was found in four families and thus the most prevalent variant in *TYR*. This variant is frequent in the South Asian population with an allele frequency of 0.0005273 (gnomAD v4.1.0) but has also been reported in the European (non-Finnish) population albeit with a much lower allele frequency of 0.00001358 (gnomAD v4.1.0). Another frequent variant was identified in *OCA2*, c.1045-15T>G (likely pathogenic); this variant has exclusively been reported in the South Asian population with an allele frequency of 0.0003953 (gnomAD v4.1.0). Nine variants were novel, and were not previously reported (**Table 1**).

**Table 1.**
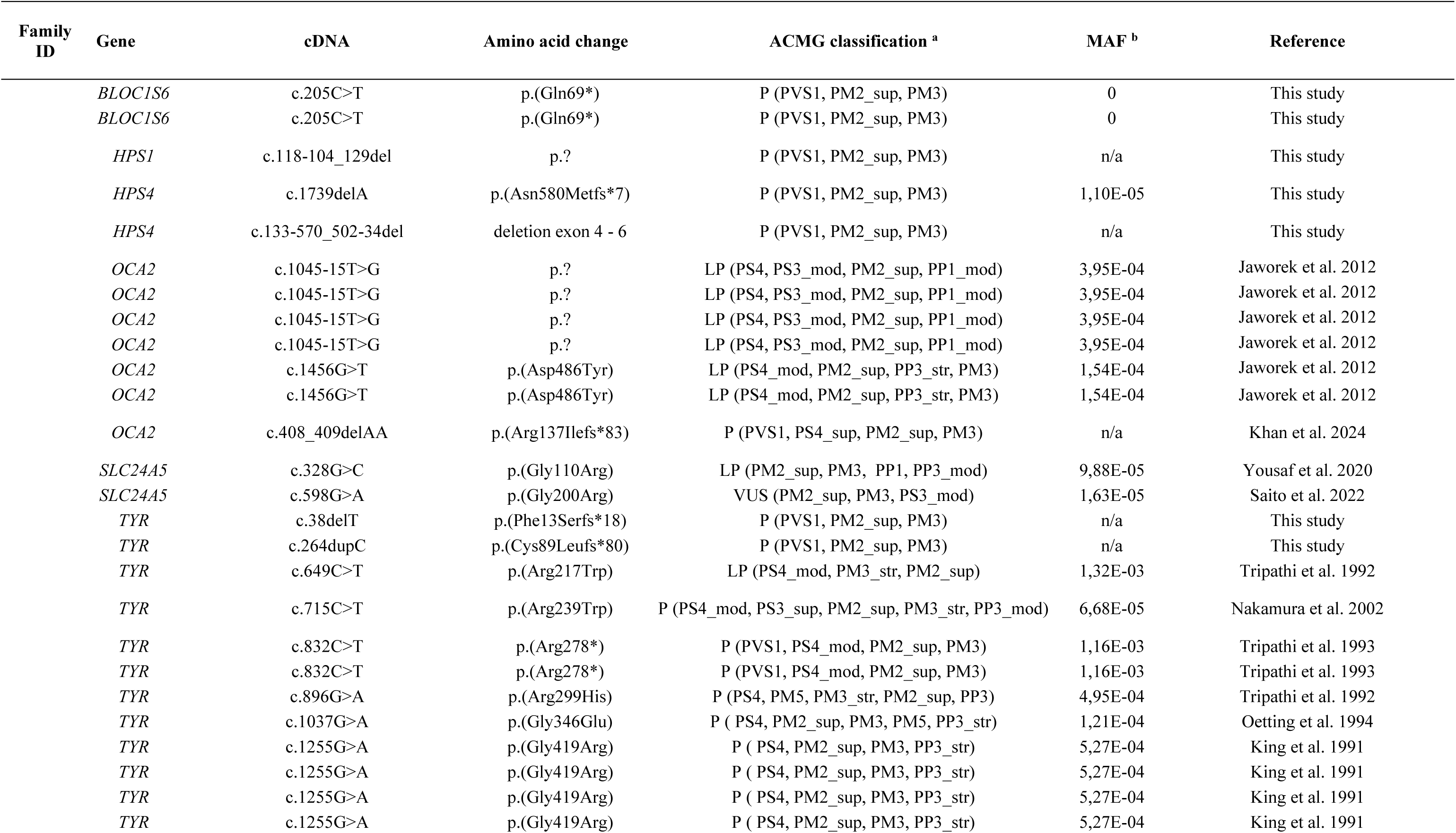
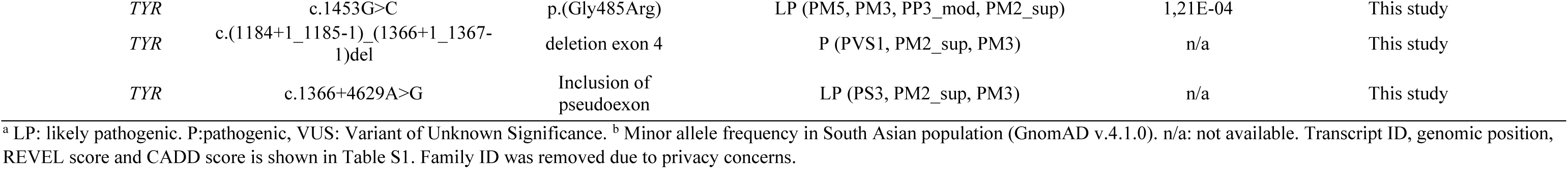
Rare homozygous variants identified in 28 consanguineous OCA families.

| Family ID | Gene | cDNA | Amino acid change | ACMG classification <sup>a</sup> | MAF <sup>b</sup> | Reference |
| --- | --- | --- | --- | --- | --- | --- |
|  | *BLOC1S6* | c.205C>T | p.(Gln69*) | P (PVS1, PM2_sup, PM3) | 0 | This study |
|  | *BLOC1S6* | c.205C>T | p.(Gln69*) | P (PVS1, PM2_sup, PM3) | 0 | This study |
|  | *HPS1* | c.118-104_129del | p.? | P (PVS1, PM2_sup, PM3) | n/a | This study |
|  | *HPS4* | c.1739delA | p.(Asn580Metfs*7) | P (PVS1, PM2_sup, PM3) | 1,10E-05 | This study |
|  | *HPS4* | c.133-570_502-34del | deletion exon 4 - 6 | P (PVS1, PM2_sup, PM3) | n/a | This study |
|  | *OCA2* | c.1045-15T>G | p.? | LP (PS4, PS3_mod, PM2_sup, PP1_mod) | 3,95E-04 | Jaworek et al. 2012 |
|  | *OCA2* | c.1045-15T>G | p.? | LP (PS4, PS3_mod, PM2_sup, PP1_mod) | 3,95E-04 | Jaworek et al. 2012 |
|  | *OCA2* | c.1045-15T>G | p.? | LP (PS4, PS3_mod, PM2_sup, PP1_mod) | 3,95E-04 | Jaworek et al. 2012 |
|  | *OCA2* | c.1045-15T>G | p.? | LP (PS4, PS3_mod, PM2_sup, PP1_mod) | 3,95E-04 | Jaworek et al. 2012 |
|  | *OCA2* | c.1456G>T | p.(Asp486Tyr) | LP (PS4_mod, PM2_sup, PP3_str, PM3) | 1,54E-04 | Jaworek et al. 2012 |
|  | *OCA2* | c.1456G>T | p.(Asp486Tyr) | LP (PS4_mod, PM2_sup, PP3_str, PM3) | 1,54E-04 | Jaworek et al. 2012 |
|  | *OCA2* | c.408_409delAA | p.(Arg137Ilefs*83) | P (PVS1, PS4_sup, PM2_sup, PM3) | n/a | Khan et al. 2024 |
|  | *SLC24A5* | c.328G>C | p.(Gly110Arg) | LP (PM2_sup, PM3, PP1, PP3_mod) | 9,88E-05 | Yousaf et al. 2020 |
|  | *SLC24A5* | c.598G>A | p.(Gly200Arg) | VUS (PM2_sup, PM3, PS3_mod) | 1,63E-05 | Saito et al. 2022 |
|  | *TYR* | c.38delT | p.(Phe13Serfs*18) | P (PVS1, PM2_sup, PM3) | n/a | This study |
|  | *TYR* | c.264dupC | p.(Cys89Leufs*80) | P (PVS1, PM2_sup, PM3) | n/a | This study |
|  | *TYR* | c.649C>T | p.(Arg217Trp) | LP (PS4_mod, PM3_str, PM2_sup) | 1,32E-03 | Tripathi et al. 1992 |
|  | *TYR* | c.715C>T | p.(Arg239Trp) | P (PS4_mod, PS3_sup, PM2_sup, PM3_str, PP3_mod) | 6,68E-05 | Nakamura et al. 2002 |
|  | *TYR* | c.832C>T | p.(Arg278*) | P (PVS1, PS4_mod, PM2_sup, PM3) | 1,16E-03 | Tripathi et al. 1993 |
|  | *TYR* | c.832C>T | p.(Arg278*) | P (PVS1, PS4_mod, PM2_sup, PM3) | 1,16E-03 | Tripathi et al. 1993 |
|  | *TYR* | c.896G>A | p.(Arg299His) | P (PS4, PM5, PM3_str, PM2_sup, PP3) | 4,95E-04 | Tripathi et al. 1992 |
|  | *TYR* | c.1037G>A | p.(Gly346Glu) | P ( PS4, PM2_sup, PM3, PM5, PP3_str) | 1,21E-04 | Oetting et al. 1994 |
|  | *TYR* | c.1255G>A | p.(Gly419Arg) | P ( PS4, PM2_sup, PM3, PP3_str) | 5,27E-04 | King et al. 1991 |
|  | *TYR* | c.1255G>A | p.(Gly419Arg) | P ( PS4, PM2_sup, PM3, PP3_str) | 5,27E-04 | King et al. 1991 |
|  | *TYR* | c.1255G>A | p.(Gly419Arg) | P ( PS4, PM2_sup, PM3, PP3_str) | 5,27E-04 | King et al. 1991 |
|  | *TYR* | c.1255G>A | p.(Gly419Arg) | P ( PS4, PM2_sup, PM3, PP3_str) | 5,27E-04 | King et al. 1991 |

|  |  |  |  |  |  |
| --- | --- | --- | --- | --- | --- |
| *TYR* | c.1453G>C | p.(Gly485Arg) | LP (PM5, PM3, PP3_mod, PM2_sup) | 1,21E-04 | This study |
| *TYR* | c.(1184+1_1185-1)_(1366+1_1367-1)del | deletion exon 4 | P (PVS1, PM2_sup, PM3) | n/a | This study |
| *TYR* | c.1366+4629A>G | Inclusion of pseudoexon | LP (PS3, PM2_sup, PM3) | n/a | This study |

<sup>a</sup> LP: likely pathogenic. P:pathogenic, VUS: Variant of Unknown Significance. <sup>b</sup> Minor allele frequency in South Asian population (GnomAD v.4.1.0). n/a: not available. Transcript ID, genomic position, REVEL score and CADD score is shown in Table S1. Family ID was removed due to privacy concerns.

Five of the 28 families (18%) were found to have causative variants in genes associated with HPS, two families with pathogenic variants in *BLOC1S6* (HPS9), two families with pathogenic variants in *HPS4* (HPS4) and one family with a pathogenic variant in *HPS1*. Four of the nine novel variants were in genes known to be associated with HPS.

Two deletions encompassing one or more exons were found, one in *TYR* and one in *HPS4*, highlighting the importance of CNV analysis in molecular genetic diagnostics.

### Identification and functional analysis of a deep intron variant in *TYR*

Gene panel analysis revealed no explanation for OCA in one family (**Fig.1**). Four affected siblings and their parents were whole genome sequenced. We filtered the data for rare (minor allele frequency, MAF < 0.01 in gnomAD v2.1.1 group South Asian) homozygous coding variants shared between individuals IV.1, IV.2, IV.6 and V.1. Our analysis identified one missense variant, shared by all four OCA individuals: p.Ala135Thr (NM_199290.3 c.403G>A) in *NACA2*. This variant has a MAF of 0.001581 in the South Asian population (GnomAD v. 4.1), a CADD score of 20.6 and a REVEL score of 0.353. ACMG variant classification scored the variant as VUS. *NACA2* is involved in skin inflammatory response (Hradetzky et al. 2013).

**Figure 1.**
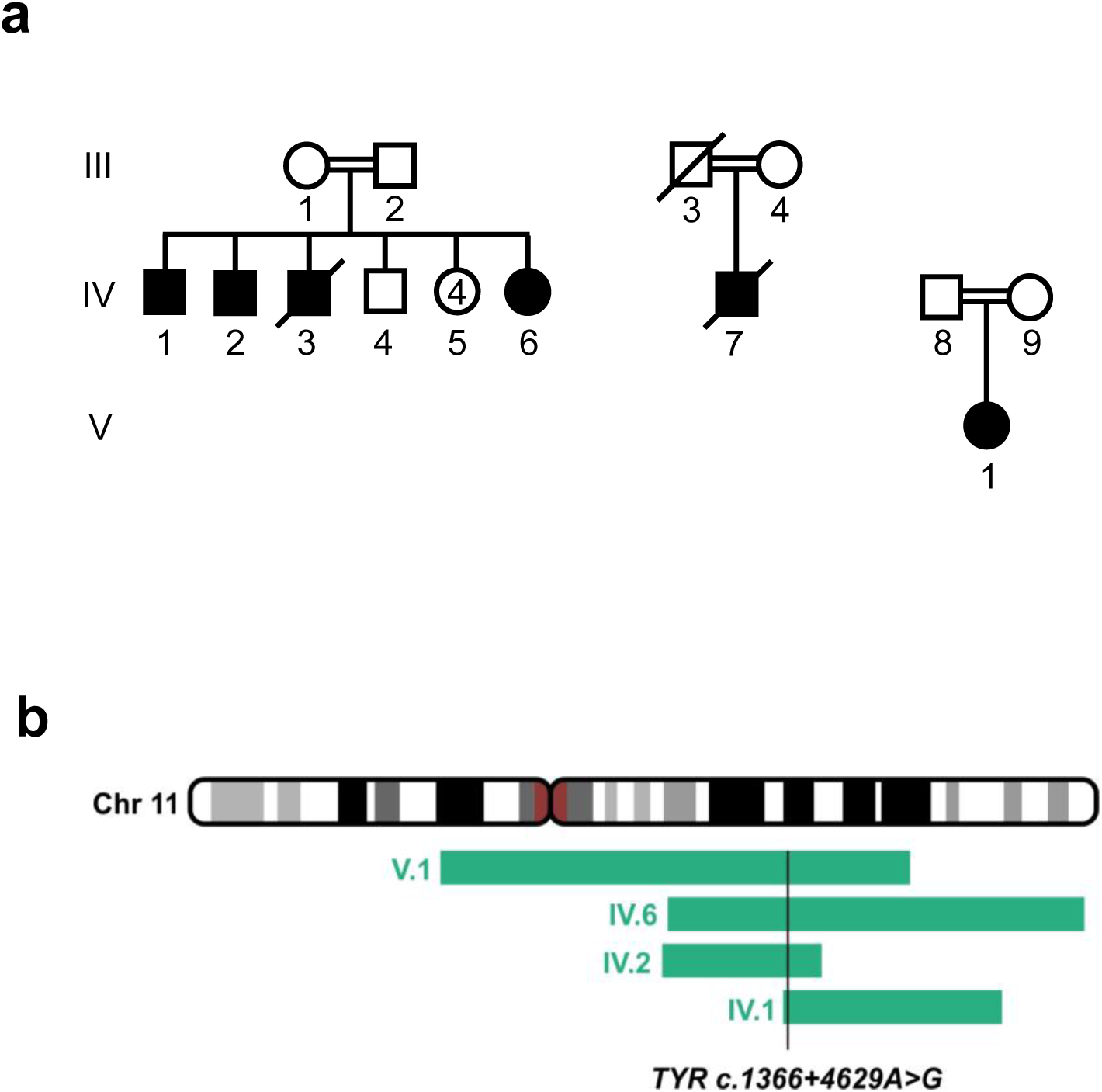
Identification and functional investigation of a pathogenic deep intron variant in TYR. a. Pedigree with six individuals presenting with OCA. b. Overlapping HBD regions between OCA individuals IV.1, IV.2, IV.6 and V.1. A shared HDB region of 5.87 Mbp contained a rare deep intron variant variant in *TYR* (c.1366+4629A>G).

Analysis of shared homozygosity by descent (HBD) regions between IV.1, IV.2, IV.6 and V.1 showed only two overlapping regions: chr11:88561408-94434662 (5.87 Mbp, which include the *TYR* gene) and chr17:55342834-63556294 (8.21 Mbp, which include the *NACA2* gene) (**Fig. S2, Fig. 1**). Because *TYR* was located in the overlapping HBD region on chr11, we analyzed both the coding and non-coding sequence of *TYR*. Filtering using a cut off MAF in gnomAD of <0.005 and selecting for variants that were homozygous in the affected siblings and heterozygous in the parents revealed seven variants located in intron sequences (**Table S1**). One variant located deep within intron 4 (hg38, NM_000372.4:c.1366+4629A>G, chr11:89289583), was extremely rare (present in one allele out of 151.994 in the gnomAD database (version 4.1.0)). *In silico* analyses using MaxEntScan indicated strengthening of a pseudo splice site (donor score increased from 8.88 to 11.00). This variant segregated with OCA in the family with an autosomal recessive inheritance pattern. Because disease-causing pseudoexons can be spliced into the mRNA when variants create or strengthen intronic pseudo splice sites (Petersen et al. 2022) we analyzed for nearby acceptor splice sites. Interestingly, we identified a matching acceptor splice site (MaxEnt Score 4.1) located 139 nt upstream. Together these pseudo splice sites define a potential pseudoexon (PE3) of 139 nt, which is likely to be strongly activated by the increase in strength of the donor splice site caused by the NM_000372.4:c.1366+4629A>G variation. Analysis of RNA-seq data confirms that this pseudoexon is included at very low levels in samples from individuals with the weak splice donor site associated with the major allele (**Fig. S3**).

In order to investigate the effect of the NM_000372.4:c.1366+4629A>G variation, we established a splicing reporter minigene.

### Minigene analysis of a deep intron variant in *TYR*

We examined the mRNA splicing across *TYR* intron 4 in a minigene containing a size-reduced intron 4 and part of the flanking exons 4 and 5 and with either the wild type or the mutant c.1366+4629A>G sequence. Minigenes were transfected into human retinal pigment epithelial cell line (RPE-1). Minigene analysis show a complex splicing pattern of *TYR* intron 4 (**Fig. 2a**). The wild type minigene (c.1366+4628A) gives rise to the normal transcript combining exon 4 and exon 5, but also several different uncommon splicing isoforms. In the context of the c.1366+4628G mutation, there is no normal splicing between exon 4 and 5. Sanger sequencing of the bands determined the identity of the pseudoexons included. Besides the PE2 pseudoexon which is in fact an annotated exon, an upstream pseudoexon (PE1) and a downstream (PE3) pseudoexon and combinations of these were identified. The c.1366+4628G mutation is located at position +5 of the donor splice site of PE3, increasing the splice site strength by altering the non-consensus A to a G (consensus) and forcing splicing from the mutant minigene to include PE3.

**Figure 2.**
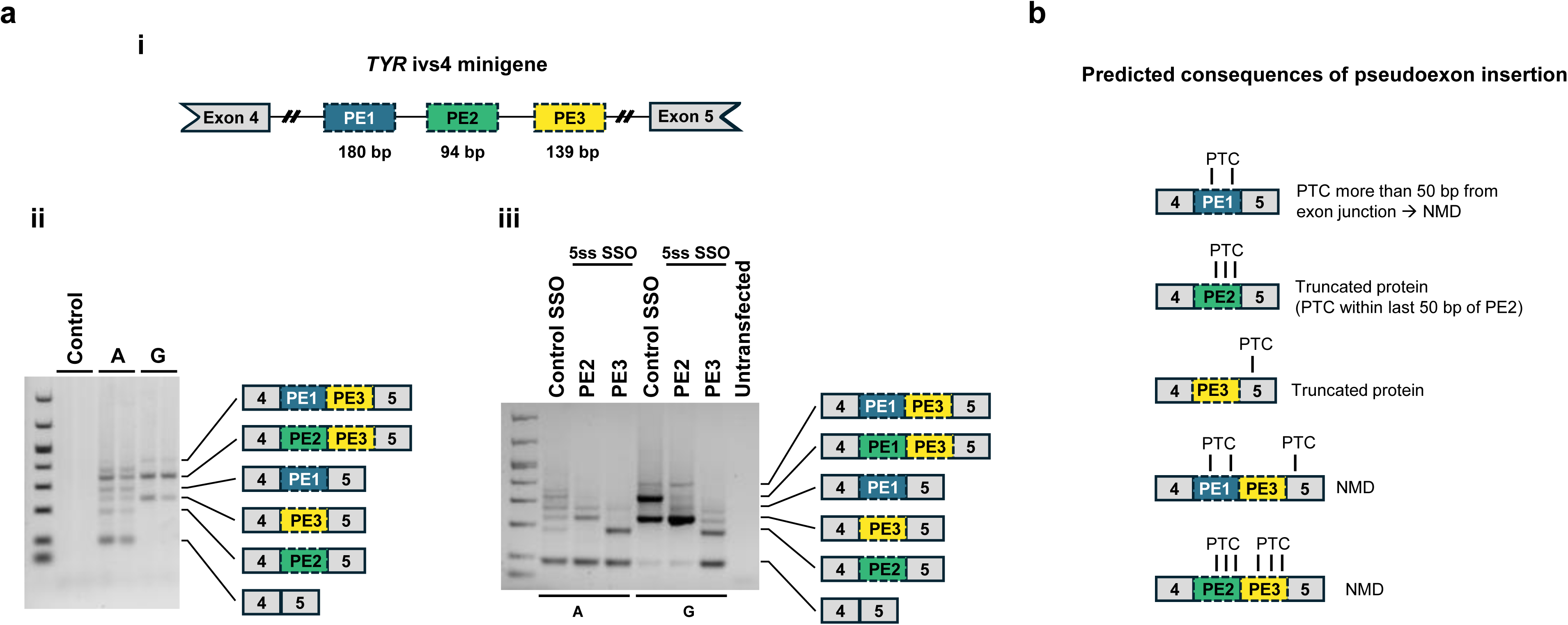
Minigene analysis of a deep intron variant in TYR. a. (i) Structure of a minigene covering the TYR exon4-exon4 region. Localization of three pseudoexons (PE1-3) are indicated, but not drawn to scale (see Fig S3 for detailed localization of the pseudoexons). (ii) RT-PCR fragments obtained from expression of minigenes containing the reference allele (A) and alternative allele (G) of the TYR c.1366+4629A>G variant identified in family with OCA (Fig.1). (iii). Results of transfection with splice switching oligonucleotides (SSOs). The sequence of pseudoexons in minigene transcripts are indicated by schematics on the right (a. ii and iii). b. Predicted consequences of pseudoexon insertion (see main text for details).

We further examined the splicing pattern of *TYR* intron 4 carrying the c.1366+4628G mutation by transfection of SSOs blocking the 5’ splice site of either PE2 or PE3. In this way, we can shift the inclusion ratio of the pseudoexons. Transfection of the PE3 5’ss SSO (and not the PE2 5’ss SSO) corrected splicing of the mutant minigene and mediated a substantial increase in the amounts of correctly spliced transcript. The introduction of PE3 either alone or in combination with PE2 or PE1 disrupts the normal reading frame; PE3 alone will introduce 46 new amino acids to the C-terminal end of the protein and cause a frame-shift in exon 5 leading to a truncated exon 5 (**Fig. 2b**). Both PE1 and PE2 introduce a PTC and therefore we expect that, *in vivo,* isoforms where PE3 is included in combination with PE1 or PE2 will be degraded by NMD. Due to the lack of a *TYR* open reading frame we expect no NMD in the transcripts from the minigene. To possibly identify additional deep-intron variants, which might lead to inclusion of PE3, we screened seven Danish OCA patients, heterozygous for pathogenic variants in *TYR*, for variants in a region of 200 bp surrounding c.1366+4629. However, this analysis did not reveal additional causative deep intron variants.

## Discussion

A molecular genetic diagnosis was identified in all analyzed families; this contrasts with studies of European populations where a detection rate of approximately 70 % are common (Chan et al. 2023; Lasseaux et al. 2018). The high diagnostic yield might be due to the inclusion criteria primarily based on hypopigmentation of the skin and hair, while in other studies, especially in cohorts from lighter pigmented populations, the inclusions criteria are often the eye symptoms. Biallelic variants in *TYR* as well as in *OCA2*, *HPS1*, *HPS4* and *BLOC1S6* causes extreme hypopigmentation of skin and hair, and variants in these genes explains OCA in all of the 28 families; in contrast, we found no variants in for example *LRMDA* which is known to cause a phenotype almost restricted to the eyes.

Several studies of molecular genetic screening of individuals with OCA from Pakistan have been performed, however, in most studies only genes associated with non-syndromic OCA were analyzed (Ullah 2022). In a total of 168 families 90 (54%) could be explained by biallelic variants in *TYR*, 59 families (35%) could be explained by biallelic variants in *OCA2* and 19 families (11%) by other genes. This corresponds well with the finding in the present study with *TYR* accounting for 54 % of families followed by *OCA2* which explained 25 % of families.

Variant c.1255G>A, p.(Gly419Arg) has previously been speculated to be a Pakistani founder mutation (Shakil et al. 2019). We identified this variant in four families, thus our data support that c.1255G>A is a founder variant. Variant c.1045-15T>G in *OCA2* was also identified in four families in our study and is exclusively found in the South Asian population. We propose that this variant might also represent a Pakistani founder mutation.

Importantly, we identified causative variants in genes associated with HPS in 18% of families. In two recent studies, eight and nine consanguineous Pakistani families, respectively, were analyzed by NGS panel which included genes for HPS or exome sequencing (Khan et al. 2024; Shakil et al. 2022). Among the seventeen families included in these studies, four different pathogenic variants were identified in *HPS1,* corresponding to 23.5%. Although numbers are small, we note that the frequency of HPS in our patient cohort is similar to previous reports, suggesting a relatively high prevalence of HPS in the Pakistani population. The main reason to perform molecular genetic diagnosis of individuals with albinism is to identify those with syndromic forms of albinism due to the required clinical care. Consequently, it should be emphasized that genes associated with HPS and CHS should be included in molecular genetic diagnostics offered to individuals with albinism.

More than expected individuals with albinism are heterozygous for pathogenic variants in *TYR*. This has led to speculations that causative variants should be found in regulatory or deep intron regions. We report for the first time a deep intron variant in *TYR* and show that the mutation c.1366+4628G disrupts the normal splicing of *TYR* exon 4 to exon 5. Instead combinations of the pseudoexons, PE1, PE2, and PE3 are included leading to PTC, truncated protein, and in many cases most likely degradation of the final mRNA transcript by NMD decreasing gene expression. Inclusion of PE3 alone leads to insertion of an incorrect amino acid sequence following exon 4 and protein truncation. Based on the crystal structure of TYRP1, which has high homology to TYR, Lai et al. (Lai et al. 2018) made a protein structure model of TYR. In this model, the transmembrane domain of TYR resides from amino acid 476 followed by the C-terminal domain harboring a melanosomal sorting signal. Since the last amino acid encoded by exon 4 is 455, we expect that if a protein is made from mRNA with PE3 inclusion this will result in a truncated protein which cannot localize correctly in the membrane of the melanosomes likely abolishing tyrosinase function.

PE2 is an annotated exon and also PE1 and PE3 can be identified in publicly available RNA seq data. This suggests that *TYR* also *in vivo* has a complex splicing pattern and that part of the mRNA could be lost due to unproductive splicing in of pseudoexons (Spangsberg Petersen et al. 2024). Because inclusion of pseudoexons typically lead to degradation by the NMD pathway, the inclusion of pseudoexons is underestimated and a rather large proportion of transcripts may be degraded due to inclusion of these pseudoexons. We show here that SSOs targeting the 5’ splice sites of PE2 and PE3 can reduce the amounts of pseudoexon inclusion and increase the amounts of correctly spliced transcript. SSO treatment that reduces unproductive splicing could therefore in principle be a way to increase gene expression of *TYR* even from alleles that do not harbor the PE3 activating c.1366+4628G variation. Such a strategy could be beneficial in cases where mutations maintain some degree of enzyme activity as previously observed in the *PCCA* gene (Spangsberg Petersen et al. 2024).

Screening of a Danish cohort of unexplained OCA patients did not reveal additional pathogenic deep intron variants in the region surrounding PE3, but nevertheless our results suggest that deep intron variants in *TYR* might be causative in unexplained OCA cases.

In summary, our data support a high frequency of HPS among OCA patients in the Pakistani population and we report the first pathogenic deep intronic variant in *TYR*.

## Supplementary information

Figure S1-S3

Table S1

## Acknowledgements

We thank the patients and their families for their participation in the project. We thank Marriam Babar for assistance with sample collection and construction of pedigrees

## Author contribution

KG, LAL and SB conceived the study. LAL, KG and BSA supervised experiments and analyses. KG, LAL, BSA and GHB wrote the manuscript with input from all other authors (MF, MVKS, LK, SB, TKD, HA, UA, IA). SB, MF, HA, UA and IA performed clinical analyses and collected pedigree information and samples. KG and LAL performed human genetic analyses. GHB, TKD and BSA performed minigene experiments and analyses. All authors critically revised and approved the final version of the manuscript.

## Data availability

Individual genome sequencing data cannot be shared due to concerns over patient privacy. Other data generated or analyzed during this study are included in the main paper, its additional files or available on request.

---

**Figure S1.** Pedigree information was removed on request from MedRxiv staff, due to privacy concerns. The pedigrees of 28 consanguineous OCA families are available from Lars Allan Larsen.

**Figure S2.**
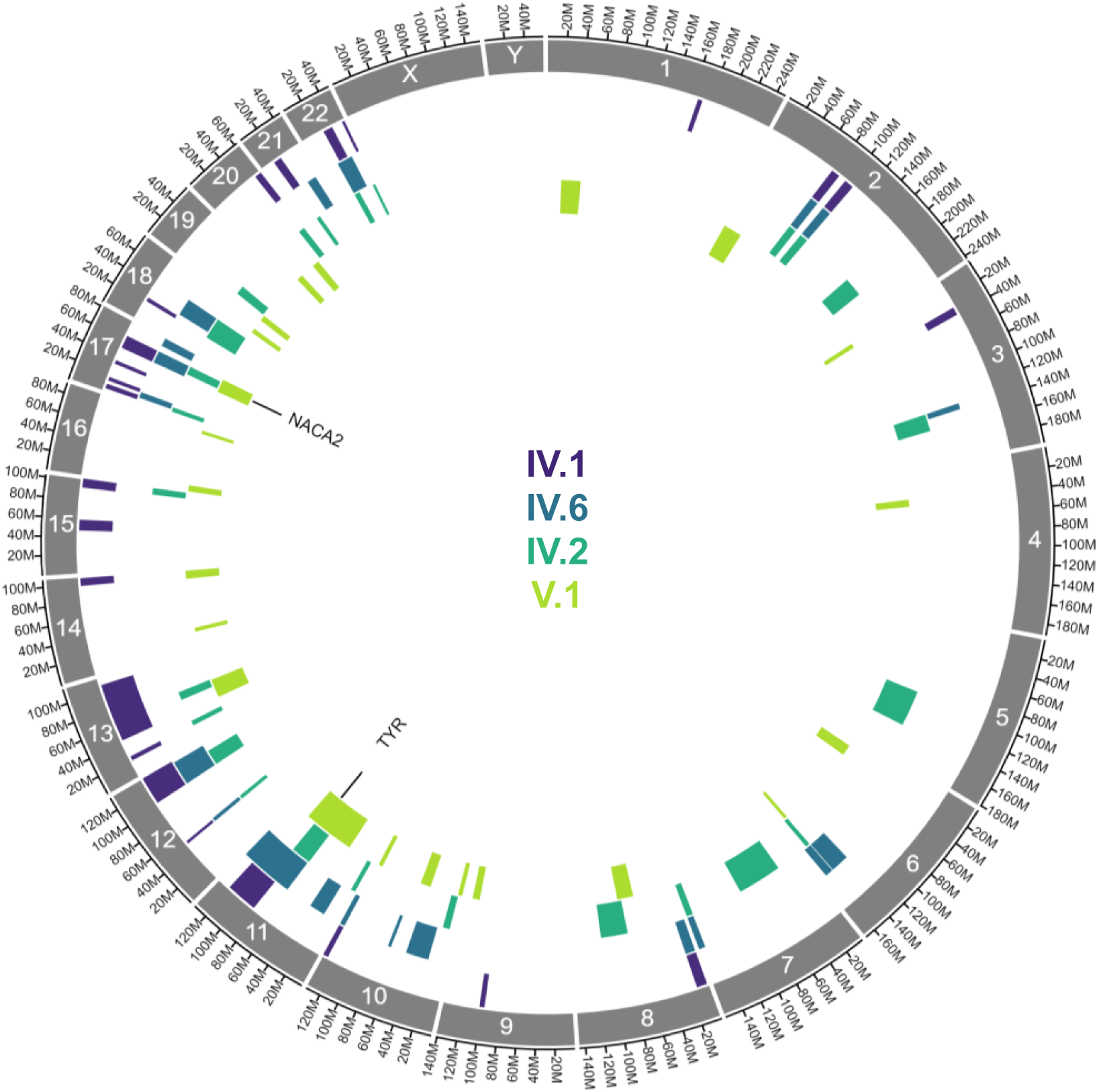
Position of homozygosity-by-descent (HBD) regions in family with OCA. HBD regions from four affected family members are indicated with colour-codes. Only regions larger than 1 mbp is shown. Candidate genes in HBD regions, shared by all four affected individuals are indicated.

**Figure S3.**
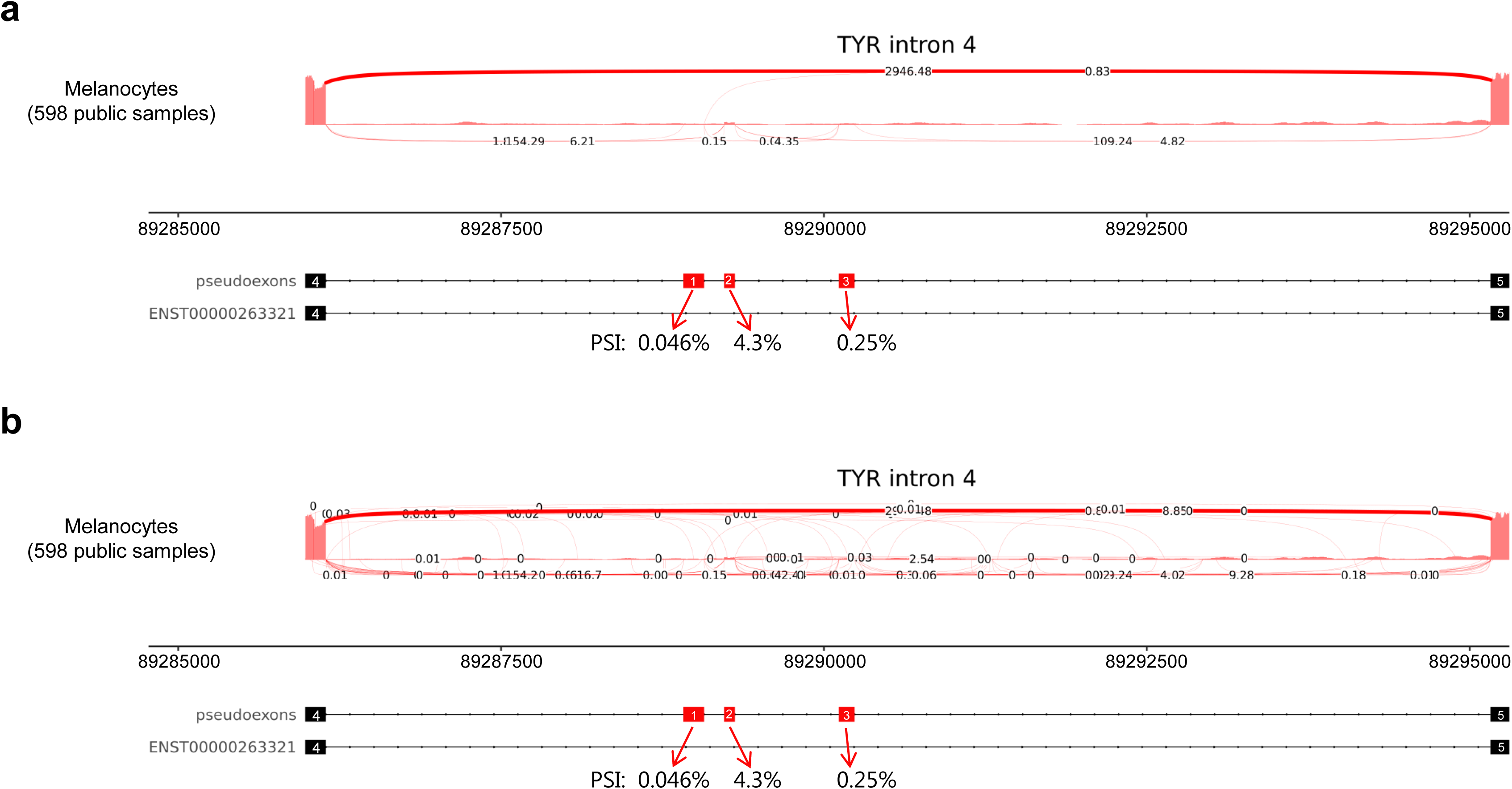
Sashimi plot of intron 4 of the *TYR* gene. Plots are based on 598 publicly available RNAseq datasets from melanocytes. The position of exon 4, Exon 5 and pseudoexons PE1-3 are indicated below each plot. Exon-inclusion ratio in TYR mRNA (percent spliced in, PSI) is shown for each pseudoexon.

